# Temporal and spatial characteristics of the spread of COVID-19 in Rio de Janeiro state and city

**DOI:** 10.1101/2020.05.13.20101113

**Authors:** Douglas S. R. Ferreira, Paula F. R. S. Ferreira, Paulo S. L. Oliveira, Jennifer Ribeiro, Elicardo A. S. Goncalves, Andrés R. R. Papa

## Abstract

From the first cases detected in Wuhan, China, of infections by the disease of the new coronavirus, COVID-19, until the present moment of this pandemic, millions of people have already been infected and hundreds of thousands have died worldwide. The way in which the virus has been dispersed in Brazil, and more specifically in Rio de Janeiro, is the motivation of the present work. Our studies consist of analyzing temporal and spatial characteristics of the spread of COVID-19 in the municipalities of the state of Rio de Janeiro and in the neighborhoods of the state capital, based on open data published by the Health Departments of Governments of the State of RJ and the Municipality of Rio de Janeiro, covering the period from February 27, 2020 to April 27, 2020. For that, we use analysis of time evolution graphs and mappings of spatial distributions and statistics analysis of spatial correlation. Our results suggest that the initial stages of spreading the virus across the state occur exponentially, with specific regions with a higher concentration of rates of cases, deaths and recovered people. In addition, our qualitative and quantitative results, for the data considered, point out that the regions with the highest income average per capita have higher rates of confirmed cases and recovered people, however, the regions of higher lethality do not are found in these places, with several of these regions of high lethality in places of low income per capita. Our results reinforce the idea of creating specific and strategic plans for policies to combat the spread of COVID-19 in the various localities in Rio de Janeiro.

## 1. Introduction

In December 2019, in Wuhan City, province of Hubei, China, several cases of pneumonia the unknown were reported. More than half of the patients with such infection has been in the *Huanan Seafood* market, a place identified as the starting point for the new coronavirus, COVID-19 [1, 2, 3, 4]. Due to its high transmission rate, which reached China and several countries in a few weeks, on March 11, 2020 the World Health Organization (WHO) stated that the new COVID-19 infection would pass to be characterized as a pandemic^1^. In Brazil, all states of the federation already report confirmed cases, of which São Paulo and Rio de Janeiro are the ones with the largest number of infections and deaths by COVID-19

Rio de Janeiro is the state with the second largest number of confirmed cases and deaths due to the COVID-19 pandemic according to data from the Ministry of Health^2^. It is worth mentioning that Rio de Janeiro is a state of great importance within the Brazilian scenario, having the second largest economy in the country and one of the most globalized cities in the world. Rio de Janeiro is an industrial hub and shelter headquarters of several multinational companies, covering food, metallurgical, steel, with emphasis on the oil extraction and refinement sectors. The tourist sector in Rio de Janeiro raises billions of Brazilian reais per year, and part of its capital was declared in 2017, as heritage of humanity by the United Nations Educational, Scientific and Cultural Organization (UNESCO). As a result of such circumstances the flow of people is high, a factor that probably contributed to the sustained transmission of COVID-19 and its rapid dissemination. Highlights also the fact that COVID-19 can also be disseminated by asymptomatic individuals, which makes it more difficult control [3, 5].

Even after the recent work by Cavalcante and Abreu [6], published on April 22, 2020, there is still a shortage of scientific studies about the epidemic evolution of the outbreak of COVID-19 in the state of Rio de Janeiro. Thus, in the present work, temporal and spatial analyzes of the dynamics of distribution of confirmed cases, recovered people, mortality rates and lethality rates due to the new coronavirus are developed. In these analyzes we take into account the places where contagions were recorded, as well as demographic characteristics like population size and per capita incomes. Thus, using statistical methods of space correlations, we developed an exploratory data analysis that are distributed in space, but that evolve over time, making it possible for us to look for association patterns between different regions and socio-economic conditions. Therefore, this study aims to contribute to a better understanding of how contagion has evolved spatially and temporally in the population of the state and the municipality of Rio de Janeiro, especially in the initial periods of its dissemination, since our studies use data that include information until April 27, 2020.

## 2. Clinical and epidemiological aspects of COVID-19

In order to make this work as self-contained as possible, we present in this section some characteristics and epidemiological information about COVID-19.

SARS-CoV-2 is a human coronavirus in the group β-coronavirus, belonging to the *Coronaviridae* family and, like other family members, it is an enveloped positive RNA virus. It was identified for the first time on January 7, 2020 [7], and it is likely that it first appeared in Malaysian pangolins (Manis *javanica*) and later passed to humans, with bats serving as a reservoir for the virus [8, 9, 10, 3].

Infection of this new coronavirus (namely, COVID-19) is triggered by the binding of the virus spike (S) protein to the converting enzyme angiotensin 2 (ACE2), which is highly expressed in the lungs and heart [11, 12]. SARS-CoV-2 invades especially epithelial cells of the alveoli, explaining why the disease is characterized mainly by respiratory symptoms [11].

COVID-19 has a fast dispersion and its transmission occurs by droplets or aerosols of saliva at the time of sneezing, coughing or talking and direct contact with saliva or respiratory system secretion of a sick person, even if asymptomatic [13, 14, 15]. The virus can still be found on contaminated surfaces, including plastics, fabrics and wood [2], which can become indirect means of contagion [13, 10]. Yet, samples of the SARS-CoV-2 genetic material can be also found in feces, thus the fecal-oral route may become a possibility of transmission and dissemination of the virus [16]. Such factors contribute to the rapid dissemination and spread of COVID-19. In view of these aspects, until the date of writing of this paper, across the planet the cases confirmed has reached 4 million people, with 270 thousand deaths.

## 3. Epidemiological and demographic data

The epidemiological data from COVID-19 for the state and the municipality of Rio de Janeiro were obtained on April 30, 2020, through the free access databases of the Health Department of the Government of the State of Rio de Janeiro^3^ and the Health Department of the Municipality of Rio de Janeiro^4^. These data cover the period of February 27, 2020 to April 27, 2020. However, we emphasize here the fact that not all municipalities and neighborhoods have data until the 27th of April, since not all city halls update their data often because each one has its peculiarities and specific difficulties.

In our study, data from confirmed cases, deaths and recovered people were used. It is worth noting that the criterion adopted by the Health Department of the State Government of Rio de Janeiro to calculate the number of recovered people considers all confirmed cases that after 14 days from the notification date or symptom onset date, are not hospitalized and did not die. In the present work we will adopt the same criteria for obtaining and analyzing our recovered data.

With a population of 15,989,929 people, according to the last census conducted in 2010 by the Brazilian Institute of Geography and Statistics – IBGE, and an estimate of 17,264,943 of in 2019 ^5^, the state of Rio de Janeiro has 92 municipalities, of which, as of April 27, 78 municipalities had at least 1 confirmed case of infection by the virus.

The municipality of Rio de Janeiro had a population of 6,320,446 people according to the 2010 census, and an estimate 6,718,903 people in 2019 ^6^, where among the 163 neighborhoods belonging to the municipality, 153 had at least 1 person proven to be infected with COVID-19 by 27 April. It is worth mentioning that, among the 92 municipalities in the state, the city of Rio de Janeiro was chosen for the study because, for the data we collect, it concentrates more than 65% of the confirmed cases of COVID-19 from across the state, as well as 60% of deaths.

Through Getúlio Vargas Fundation’s Social Policy Center, we also obtained the distribution of population average income per capita of each municipality in state^7^and of each neighborhood in the municipality of Rio de Janeiro^8^.

## 4. Methods

As previously mentioned, in the present work we perform temporal and spatial analyzes of the cases of COVID-19 in the population of Rio de Janeiro. For this, we use three different approaches, where the prospects despite different, are complementary to each other.

### 4.1 Time evolution

Using epidemiological data acquired in the municipal and state health services secretariats, we carry out the construction of graphs of time evolution of numbers of infected people, deaths (both in mortality and lethality) and recovered people. These analyzes allow observing the functional behavior of the spread of the virus as time goes by, as well as take a more detailed and meticulous look about the local differences that exist in each region, since it is expected that regions with different characteristics socio-economic conditions also have different contagions dynamics, once the behavior of people in these regions will be different. Such different behaviors include from the frequency with which people in these regions travel to other locations away from their homes, going through their type of work, and even the form of driving transport that these people use.

As reported in literature, different contagious diseases, such as measles, cachumba and rubella, spread through the population with the number of infected in the initial epidemic phases growing exponentially [17, 18, 19, 20], that is, obeying a temporal function which has the form:

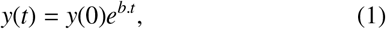

where, *y*(*t*) is the total number of the epidemiological quantity measured at time *t* and *b* is a constant that depends on the contact rate between individuals and the recovery rate (or death) of individuals.

According to previous studies using data from Brazil and the most diverse locations around the world, the dissemination of COVID-19 has a similar behavior, that is, some kind of exponential growth for the total number of infected people in the early stages of the epidemic outbreak [21, 22, 23, 24, 25, 26]. As a result, several of our time evolution graphs will be presented in *semi-log* scales, causing data that has growth exponential to be represented by an increasing straight line in this scale. In this way, our analyzes will us allow to observe how the data regarding the population of Rio de Janeiro state and city behave.

### 4.2 Spatial distribution

From the association of epidemiological and demographic data of Rio de Janeiro, we will use spatial mappings of COVID-19 distributions for confirmed cases, mortality, lethality and recoveries that occurred in the state and in the municipality of Rio de Janeiro. These analyses of spatial distribution take into account both the epidemic factor (confirmed cases, deaths or recoveries), and the social-demographic factor (population and per capita income of each municipality or neighborhood). This method seeks to perform a visual, qualitative and quantitative analysis, of how the outbreak of COVID-19 behaves through the spatial and social organization of the fluminense population, that is, this method has the objective of studying, for the outbreak of COVID-19, possible existing relationships between different locations and different population distributions and incomes.

### 4.3 Statistical analysis of spatial correlation

One scientific method of wide applicability in the most various areas of knowledge, is the statistical analysis of data. Obtaining, mining, characterizing and systematic analysis of data, based on well-defined statistical criteria, is one of the branches of science that allows us to diagnose the past, understand the present and infer possible future scenarios.

The characterization of epidemiological data disseminated in a region implies relationships between the epidemic agent and its association with the local space, where a fundamental aspect of this spatial exploratory analysis is the numerical characterization of spatial dependence. However, as expressed in [27], human perception alone, is not rigorous enough to determine patterns of significant spatial groupings. Thus, in the present work we will use a statistical method capable of assimilating these associations and extracting spatial autocorrelation using quantitative instruments and not just visual observations.

In order to quantify spatial autocorrelations between the epidemiological agent and the incidence sites, we will use *Moran’s I* statistic. This statistic provides a formal indication on whether the value observed at each point is similar or distinct from the values observed at neighboring points, that is, it measures the degree of linear association between a vector of values observed and the weighted average of the values of its neighbors [28, 29, 30].

*Moran’s I* statistic presents itself as a coefficient of spatial autocorrelation using a measure of auto-covariance in the form of a cross product, varying from −1 to 1 [31]. Mathematically, *Moran’s I* statistic can be expressed by

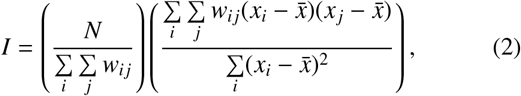

where, *N* is the number of regions considered, *x* is the value of the variable of interest, *i* and *j* are the regions, 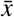 is the average of the values observed of the variable of interest and *w_ij_* are the matrix elements of spatial proximity weights (i.e., *w_ij_* is a measure of spatial proximity between regions *i* and *j*).

As a result, positive values of *I* indicate a direct correlation, negative values of *I* indicate an inverse correlation and *I*=*0* indicates no spatial correlation. Furthermore, the much the closer to 1 is the *I* module, the greater the intensity of the existing spatial correlation, and the closer to 0, the lower the intensity.

An efficient way of interpreting Moran’s *I* statistic is using the Moran dispersion diagram, where the index *I* is given by the linear regression coefficient on a graph to the value of the variable of interest in a given region of the space and the average of the values of this variable for neighbors of the considered region. With that, this diagram is divided into four quadrants, which correspond to four levels of association between the region considered and its neighbors. The quadrants I, II, III and IV, respectively, represent regions *high-high, low-high, low-low* e *high-low*.

However, when analyzing a spatial distribution in a map, it is important be able to detect local clusters regions with similar characteristics, that is, detecting significant clusters in regions of the map. For this, it is important to carry out local measures, which in this case means using Moran’s *I* statistic, defined by:

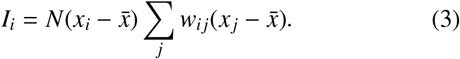

In the present work we will use the calculations of the local values of Moran’s *I* to carry out the local indicators of spatial association (LISA) [28].

It is worth mentioning that *Moran’s I* statistic was also used as a tool in other previous works on analysis of spatial correlations in the spread of diseases, such as dengue and tuberculosis, in the state of Rio de Janeiro [32, 33, 34].

## 5. Results and discussion

In accordance with the previous section, we will carry out spatio-temporal analysis of COVID-19 data in Rio de Janeiro from the perspective of the three methods presented.

It is important to highlight here the gap between the day of data collection and the time that they represent. Due to several problems, for example, time for database updates and time for examination results, the data presented in the catalogs always refer to a past time and not to the present time in which they were extracted. This means that our data, although they were obtained on April 30, 2020, actually refer to a previous time, a time that is not necessarily the same for all municipalities, since each Municipal Health Department has its own problems and therefore different lags. Yet, such difficulties do not preclude the present study since, even if there are lags in the disclosure time of data, the behavior is the same.

### 5.1 Time evolution

For the analysis of the temporal evolution of the spread of the disease the data from COVID-19 and the data on demographics of the population of Rio de Janeiro, were used. In order to minimize possible problems related to daily records epidemiological data, the analyzes presented in this work are calculated based on accumulated data and are not daily data only.

In order to carry out analyzes at different levels of spatial detail, data were analyzed for three levels of regional scales: the state as a whole, municipalities of the state, the neighborhoods of the municipality of Rio de Janeiro. The results are presented below.

#### 5.1.1 Analyzes for the state of Rio de Janeiro

Considering the entire state, Fig. 1 presents the evolution of confirmed cases, mortality, lethality and recovered people across the state of Rio de Janeiro (RJ). These graphs are constructed based on the first confirmed case and the first death in the state, occurred, respectively, on February 27 and March 12, 2020, according to the open data information from the Health Department of the Government of the State of Rio de Janeiro.

**Figure 1:**
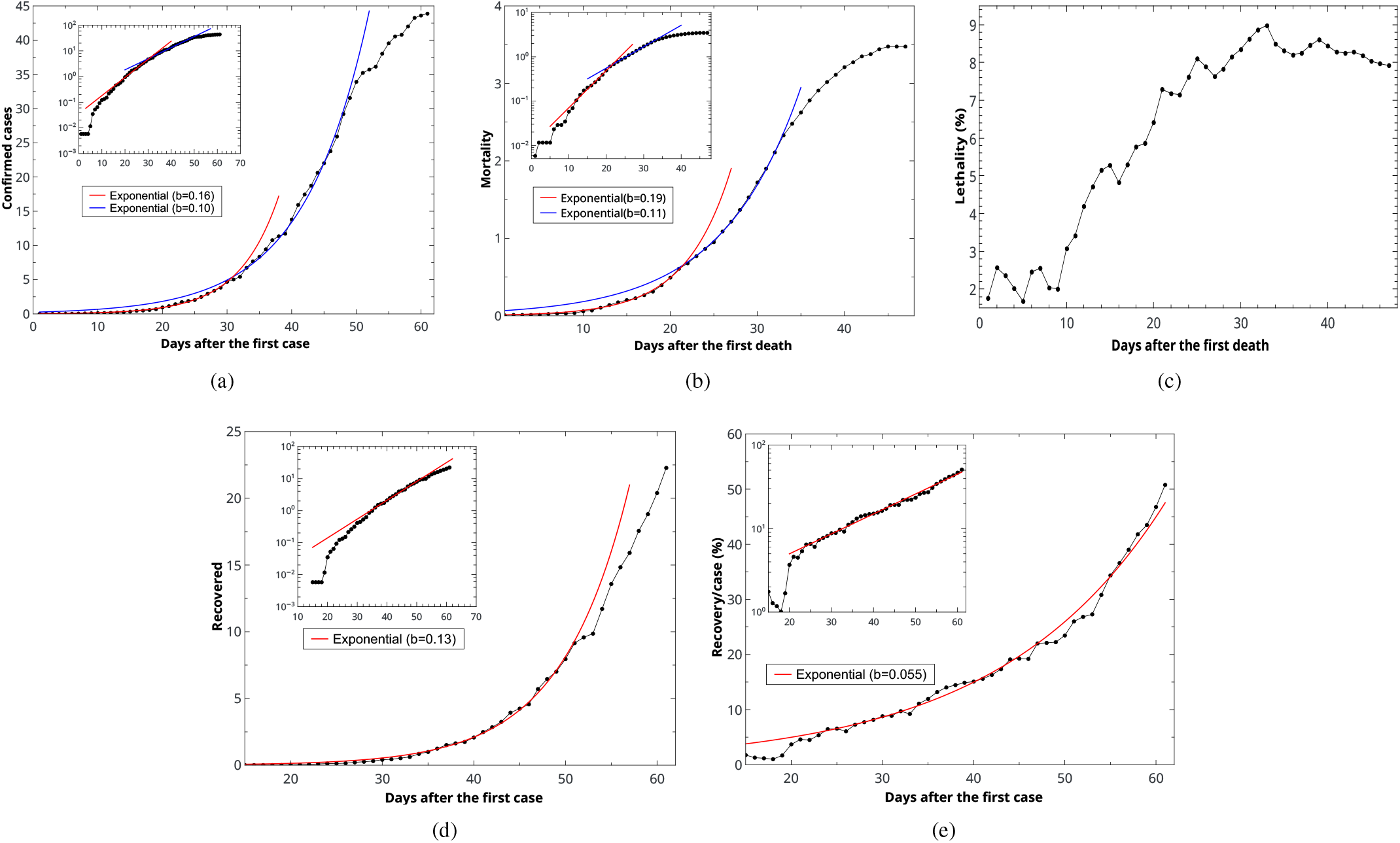
Time evolution for accumulated data of (a) confirmed cases, (b) mortality, (c) lethality, (d) recovered people and (e) people recovered by confirmed cases, per 100 thousand inhabitants, for the entire state of Rio de Janeiro. *Insets*: graphs (a), (b), (d) and (e) in *semi-log* scale. Adjustment values in the text body.

Figs. 1(a), 1(b), 1(d), 1(e) provide, respectively, the evolution of the rate of confirmed cases, mortality, lethality and people recovered throughout the state for every 100 thousand inhabitants. These graphs are presented in both a linear graph and in a *semi-log* one, so we can observe clearer how the data behaves, since an exponential function becomes a line in a *log-linear* graph.

In the graph of Fig. 1(a) we can see that we have two exponential periods. Until the 30th day the data obey an exponential function, whose adjustment for eq. (1) provides a coefficient *b* = 0.16 ± 0.01, with coefficient of determination *R*^2^ = 0.993345. Between the 30th and the 50th days of the chart the best adjustment occurs for *b* = 0.10 ± 0.01, with *R*^2^ = 0.995657.

For the mortality graph (Fig. 1(b)) two exponential behaviors arise, the first for an interval before the 20th day and the second for an interval after the 20th day. The coefficient *b* adjustments for these two intervals are, respectively, 0.19 ± 0.01 with *R*^2^ = 0.991273 and 0.11 ± 0.01 with *R*^2^ = 0.999107.

Fig. 1(d) shows the rate of people recovered for each 100 thousand inhabitants of the state. Adjusting the exponential functionl (1) to data it is obtained a value of *b* = 0.13 ± 0.02 with coefficient *R*^2^ = 0.996678. By the figure *inset*, in *log-linear* scale, it is observed that the exponential function is better adjusted for the interval between days 35 and 52.

In Fig. 1(e) we have the curve for the time evolution of the percentage recovered in relation to confirmed cases, or (*recovered*)/(*confirmed cases*). It can be seen that, for the state as a whole, this percentage grows significantly exponentially, from the 20th day until the end of the considered data. The best fit is obtained for a coefficient *b* = 0.055±0.001 with *R*^2^ = 0.987921.

It should also be noted that, although the graphics do not maintain their exponential behavior until the end of the data, this change does not necessarily indicate that exponential behavior ceased to exist in that interval of time, because despite the points no longer follow the exponential function, there is a lag in collecting and releasing the data, making the data to be corrected only a few days after the release, and with that, the last days of the catalogs lack data, producing a greater incompleteness of data at the end of the time interval considered.

In Fig 1(c) we have the evolution of the lethality rate of the virus in the state of Rio de Janeiro. We observe that after the 30th day of the first death in the state, lethality reaches a plateau approximately constant, with a value just above 8%. It is worth noting here that in the present work all calculations of lethality rates are carried out in accordance with the method adopted by the Brazilian Ministry of Health^9^, where the rate lethality of people by COVID-19 is measured by the ratio (deaths) / (total confirmed cases), instead of considering the ratio (deaths) / ratio (total completed cases). This measure was adopted to make possible to compare our results with those of different locations from the country.

However, although the graphics contained in Fig. 1 include throughout the state, they have the limitation of not being able to identify the characteristics due to the particularities existing between different regions of the state, and for that reason they will be presented later in analyzes of smaller regions, divided by municipalities and also by neighborhoods.

#### 5.1.2 Analyzes for the municipalities of the state of Rio de Janeiro

In order to produce a greater spatial detail of the dissemination phenomenon of the new coronavirus within the Rio de Janeiro state, we will present the analysis of confirmed cases, mortality, lethality and people recovered for the municipalities in the state. An observation to be made is that for the analysis of municipalities it will not be presented errors related to parameter *b* of eq. 1 nor the coefficients of determination *R*^2^. This is because all analyzes are made for large numbers of municipalities, and therefore it would not make sense to place the adjustment values for all cases. In this way the *b* values presented are the measures of the angular coefficients of straight lines shown in the *inset* graphics.

*\* Confirmed cases*: Fig. 2 shows the evolution of cases confirmed for the municipalities of Rio de Janeiro. In this figure results are presented for the 20 municipalities with highest number of cases for every 100 thousand inhabitants. With the objective of observing, the growth trend of confirmed cases in each municipality more clearly, we also used the *semi-log*, scale, where it can be noted that several municipalities have similar trends, characterized by the line present in the *inset* of Fig 2. Thus, despite the municipalities don’t behave in exactly the same way, we can observe the presence of the exponential form, represented by eq. 1, in the growth behavior of the number of confirmed cases in the municipalities of the state of Rio de Janeiro. By the *inset*, with *log-lin* axis, in the figure, we are able to fit a straight exponential that meets an average behavior of several municipalities for the interval approximately between the 20th and 40th day. This adjustment is obtained for *b* = 0.10.

**Figure 2:**
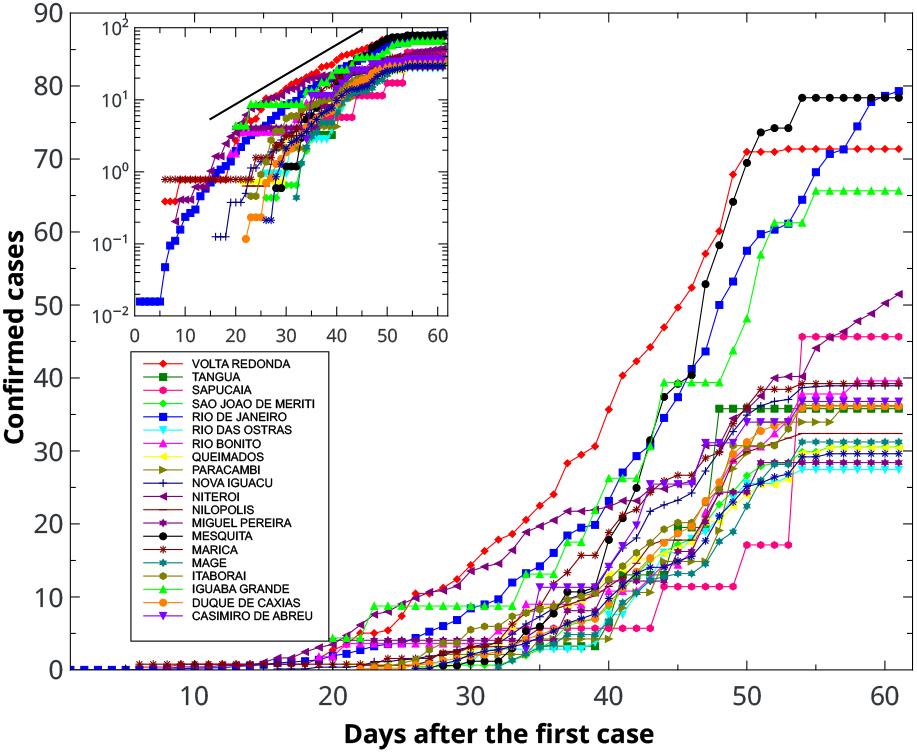
Time evolution for accumulated data for confirmed cases for municipalities in the state of RJ, after the first confirmed case of infection by COVID-19 in the state. The 20 municipalities with the highest confirmed case rates per 100 thousand inhabitants. *Inset*: *semi-log* plot. Adjustment values in the text body.

As noted in section 5.1.1, despite the graphs of the *inset* in Fig 2 have a change in behavior beginning between 40 and 50 days, this change does not indicate necessarily that exponential behavior has left to exist in that time interval, because although the points no longer follow the same behavior of the crescent line, the lag in the collection and release of data, results in data incomplete which are corrected only later.

*\* Deaths*: To analyze the spatial distribution of the evolution of deaths due to the new coronavirus, the rates of mortality and lethality by the virus in the populations of each municipality of the state were calculated. Fig. 3 shows the results for the 10 municipalities with the highest death rates. It is worth mentioning that in our calculations, only municipalities with more than 6 confirmed deaths were considered. Such measure was adopted in order to minimize statistical problems, since populations that are small compared to the average may have high mortality and lethality rates, although, for example, have 1 or 2 deaths.

**Figure 3:**
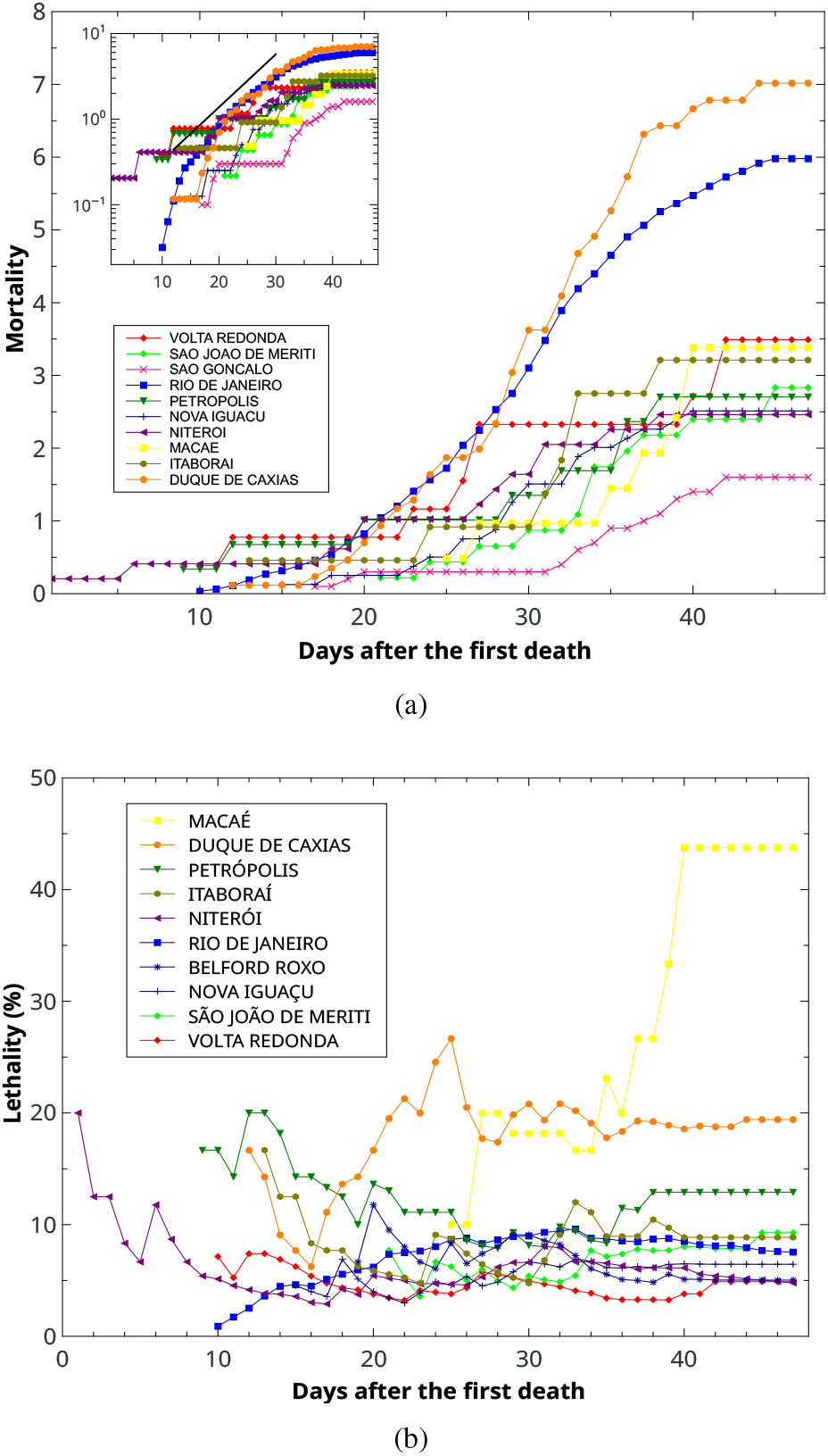
Time evolution for accumulated data on (a) mortality and (b) lethality, for the municipalities of the state of RJ, since the first death by COVID-19 confirmed in the state occured. The 10 municipalities with higher mortality and lethality values, respectively, are represented. For the calculations, only the municipalities that have more than 6 deaths in the total were considered. *Inset*: graph (a) in *semi-log* scale. Adjustment value, in the text body.

Fig. 3(a) shows the time evolution of mortality in the municipalities, that is, the number of deaths per every 100,000 inhabitants in each municipality. It can be noted that in this case the exponential behavior is present for the interval between the 15th and the 30th days (approximately), given that only in this interval we have the adjustment of a straight to the *semi-log* graph. The best adjusted value for the coefficient *b* is 0.14. In addition, it is observed that two municipalities stand out in relation to the others, being Duque de Caxias and Rio de Janeiro. Duque de Caxias, in addition to having greater mortality, still stands out for maintaining the exponential behavior until approximately the 35th day.

The temporal evolution of the lethality rate in the municipalities is described in Fig. 3(b). In this case, we have, for each municipality, the analysis of the number of deaths in relation to the number of confirmed cases of infection, using the same calculation explained in section 5.1.1. In this way, we are able to verify that the municipalities of Macaé and Duque de Caxias stand out when compared to other municipalities, having at the end of the considered period, lethality rates equal to 43.7% and 19.4%, respectively, these values being well above the value of 8% found for the state of RJ. In addition, we observe that, after the 30th day, most municipalities start to have an approximately constant lethality, with a value between 4% and 14%.

*\* Recovered*: The rate of people recovered per 100 thousand inhabitants of each municipality is shown in Fig. 4(a). In Fig. 4(b) it is represented the fraction of people who have recovered from infection in relation to the number of people who were confirmed to be infected. In both cases, the results are for the 20 municipalities with the highest rates of recovered.

**Figure 4:**
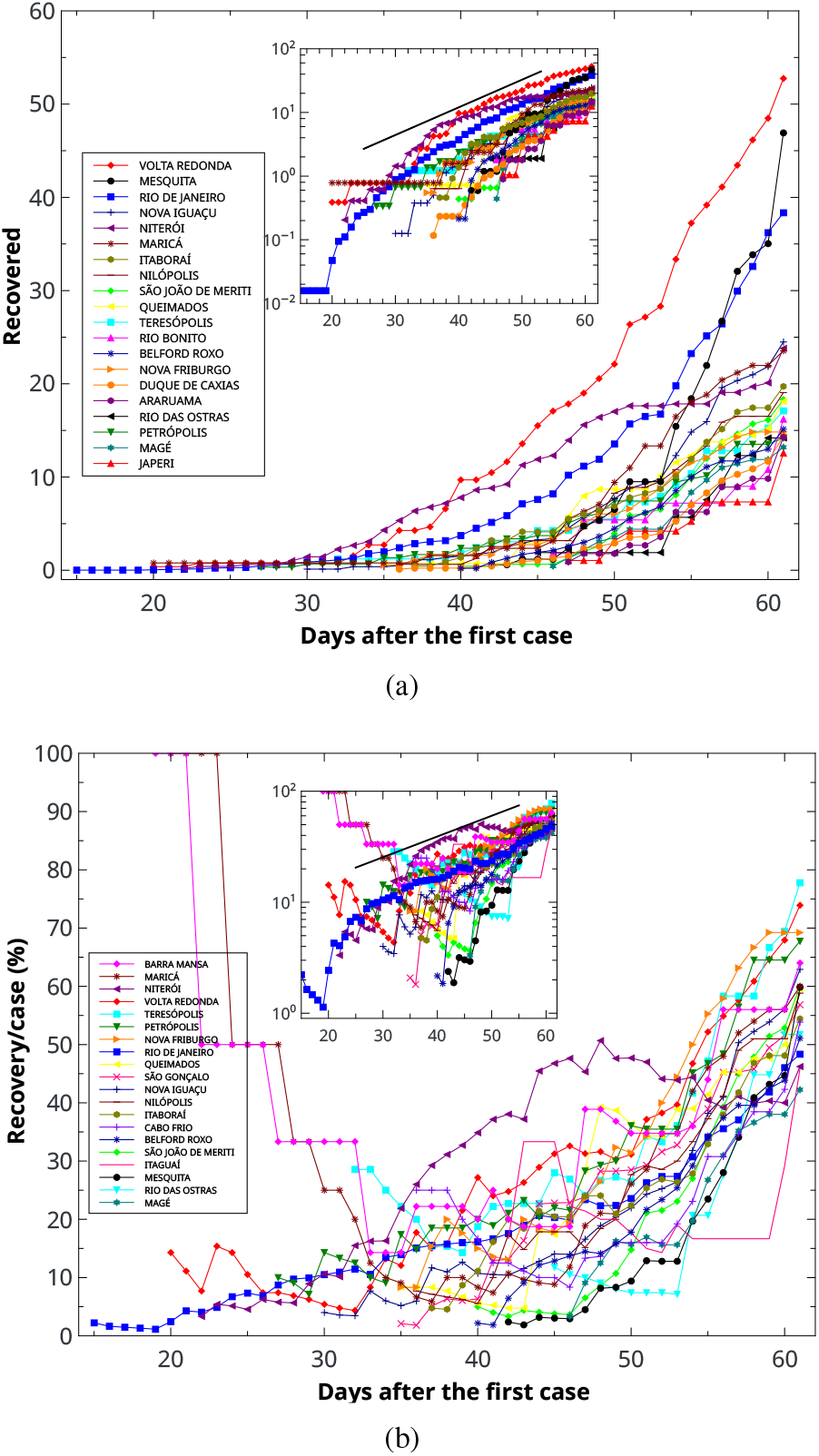
Time evolution for accumulated data of recovered people (a) per 100 thousand inhabitants and (b) by number of confirmed cases, for municipalities of the state of RJ. Both graphs represent the 20 municipalities with higher values and only the municipalities that have more than 20 confirmed cases in total. *Insets*: graphics on a *semi-log* scale. Adjustment value, in the text body.

We emphasize again that all people who have been confirmed with the virus are considered recovered and after 14

We emphasize again that there are considered as recovered all people who have been confirmed with the virus and after 14 days their clinical picture did not evolve to hospitalization or death.

A direct analysis of the graph in Fig. 4(a), shows us that the Volta Redonda municipality is highlighted in relation to others, not only for having a higher rate of people recovered, but also for having a faster growth rate of this rate. We still have that the *inset* of Fig. 4(a) indicates that the behavior of the recovery rates of some municipalities also follow an exponential, since these have the behavior of the line indicated in the figure, even if just for a short period of days. The adjustment of the line shown in figure *inset* has *b* = 0.10.

The graph of evolution of the rate of recoveries per case, Fig. 4(b), presents a scenario where the variations between the municipalities are very large and do not have a regularity pattern. However, we can observe that the behavior of data from the municipality of Rio de Janeiro have great similarity with Fig. 1(e). Thus, it is observed that the municipality of Rio de Janeiro has great influence on the behavior of this graph for the entire state. The exponential adjustment in the *inset* of Fig. 4(b) has a coefficient *b* = 0.043.

Another result observed in Fig. 4(b) is that, between the days 35 and 50, Niterói has a recovery rate per case above other municipalities.

#### 5.1.3. Analyzes for neighborhoods in the municipality of Rio de Janeiro

In order to achieve a regional investigation even more detailed, we will present below the results for neighborhoods in the municipality of Rio de Janeiro. However, we emphasize that for the neighborhoods it was not possible to carry out the analyzes of temporal evolution of deaths due to COVID-19, as the free access data made available on the Municipal Health Department website does not have information about the dates of deaths.

*\* Confirmed cases*: Similar to what was done for the municipalities of RJ, we have in Fig. 5 the number of confirmed cases per 100 thousand inhabitants for the neighborhoods of municipality of Rio de Janeiro. This figure shows the results for the 20 neighborhoods with the highest rates of confirmed cases. It is observed that the neighborhoods located in the southern part of the city have higher rates of confirmed cases. The only exception is the Bonsucesso neighborhood, which after the 40th day showed an abrupt growth greater than all other neighborhoods. The exponential behavior presented in the *inset* is better followed by the neighborhoods in the southern zone, with adjustment for *b* = 0.051.

**Figure 5:**
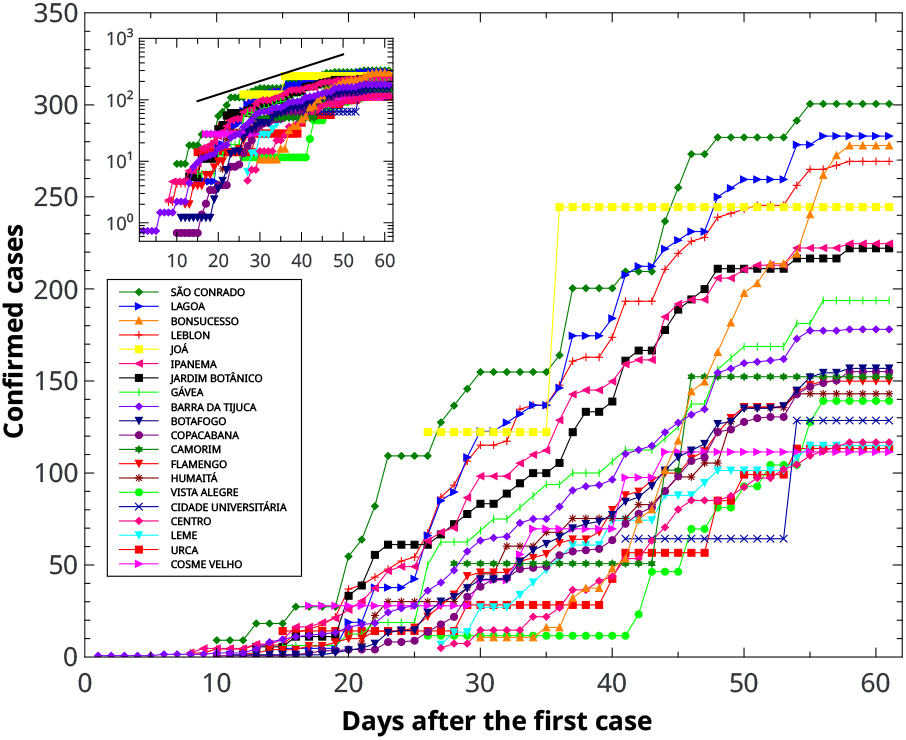
Time evolution for accumulated data from confirmed cases for neighborhoods in the municipality of Rio de Janeiro, per 100 thousand inhabitants. The 20 neighborhoods with the highest rates of confirmed cases are presented. *Inset: semi-log* plot. Adjustment values in the text body.

*\* Recovered*: Results for the rate of recovered people per 100 thousand inhabitants of the neighborhoods of Rio de Janeiro and the recovery rate in relation to confirmed cases, are shown in the graphs of Fig. 6. In both cases data are displayed for the 20 neighborhoods with the highest recovery rates. The neighborhoods with the highest recovery rates are in the southern of the municipality of Rio de Janeiro.

**Figure 6:**
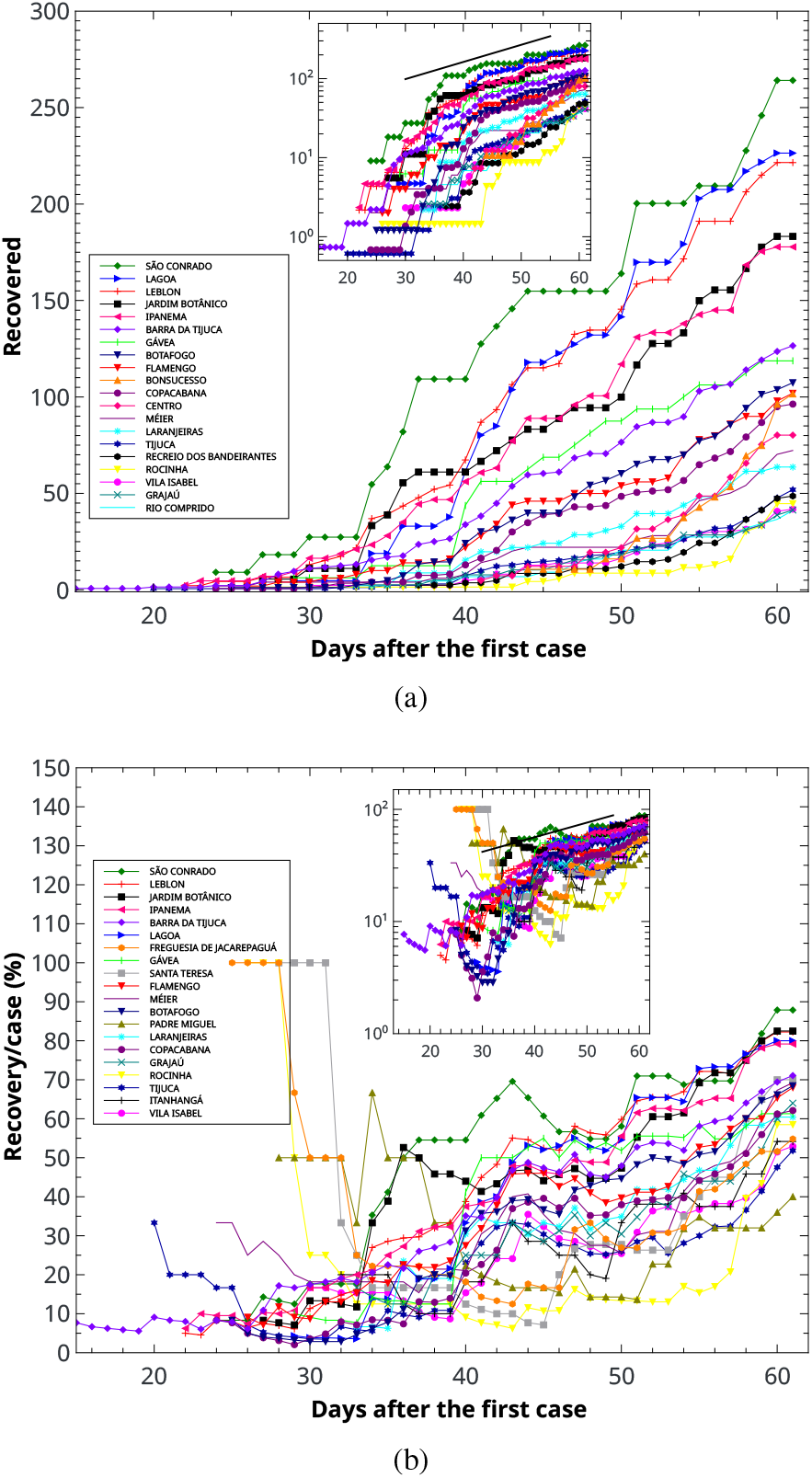
Time evolution for accumulated data of recovered people (a) per 100 thousand inhabitants and (b) by number of confirmed cases, for the neighborhoods of the municipality of Rio de Janeiro. The 20 neighborhoods with higher values, are represented. For the calculations, only the neighborhoods that have more than 20 confirmed cases in total were considered. *Insets*: graphics on a *semi-log* scale. Adjustment value, in the text body.

The *inset* in Fig. 6(a) allows us to observe the exponential expansion behavior of several neighborhoods, with an adjustment *b* = 0.050.

In Fig. 6(b), despite the irregular and heterogeneous behavior between neighborhoods, it is also possible to notice a pattern for the interval after day 35. The line adjusted in the *inset* has *b* = 0.031, indicating that for this interval the exponential behavior is more modest than the for the others.

For neighborhoods in the municipality of Rio de Janeiro, we considered recovered all persons who have been confirmed with the virus and after 14 days were not listed as hospitalized or as a death.

### 5.2 Spatial distribution

In accordance with the method described in section 4.2, we will present here a spatial distribution of epidemic data together with social-demographic data for the municipalities of RJ, as well as neighborhoods in the municipality of Rio. To achieve this objective, we will use each of the measures of epidemic rates in conjunction with the data obtained of per capita income values for each municipality and neighborhood used.

Such analyzes will be made as explained below. Cumulative data number of confirmed cases, mortality, lethality and recovered people, will be overlaid on heat maps of per capita income distributions, which will be built on geographic maps of the considered regions. All maps presented in this section are maps of spatial distribution for accumulated data, where the 10 municipalities with the highest rates per 100 thousand inhabitants are highlighted and named. In all cases the sizes of the circles in red grow according to the rate values and the heat map varies from white to blue according to the local average per capita income considered. The maps presented in this section were built using the QGIS software^10^.

It is worth mentioning here that spatial distribution maps, in general, have more data than time evolution graphs. This is due to the fact that not all data available in the databases of the Health Secretariats, have the date of occurrence. In this way it is possible to use these undated data for the construction of the maps, however, it is not possible to use them for time plots.

#### 5.2.1. Maps for the municipalities of Rio de Janeiro

*\* Confirmed cases*: In Fig. 7 we have the distribution of cases confirmed by 100 thousand inhabitants for each municipality in the state. The map shows the 10 municipalities with the highest confirmed cases indexes, where we can see that the municipality of Rio de Janeiro does not occupies the first position, even though it has a much larger population than all other municipalities. It should also be noted that the municipalities of Iguaba Grande and Sapucaia despite having only 15 and 9 cases confirmed until April 22, 2020 (where these data were obtained on April 30), respectively, occupy the fourth and seventh positions respectively. This is due to their low populations.

**Figure 7:**
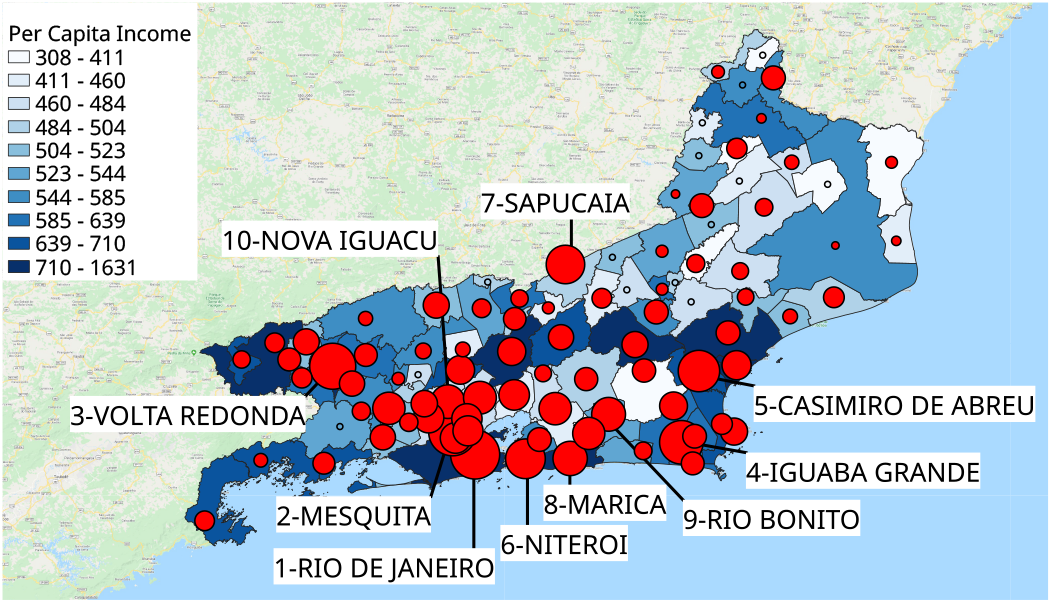
Confirmed cases for municipalities in the state of RJ. The empty circles represent locations with no confirmed cases. There are highlighted in map the 10 municipalities with the highest rates of confirmed cases in the state.

It can also be observed that most concentrations of cases occur in the vicinity of the municipalities of greater per capita income. In the same way, it is noted that the northern part of the state, when compared to the rest of the state, has a low rate of confirmed cases in its population.

*\* Deaths*: As with the time evolution graphs, the analysis of deaths is divided into two calculations. An observation to be done is that in order to avoid statistical anomalies in the results, only the municipalities which registered more than 6 deaths in the considered time interval, were considered.

Fig. 8(a) shows the spatial distribution of the rate of mortality for the municipalities, where the 10 municipalities with the highest rates are highlighted. In line with what was observed in section 5.1.2 the municipality of Duque de Caxias has the highest mortality rate in the state of Rio de Janeiro.

**Figure 8:**
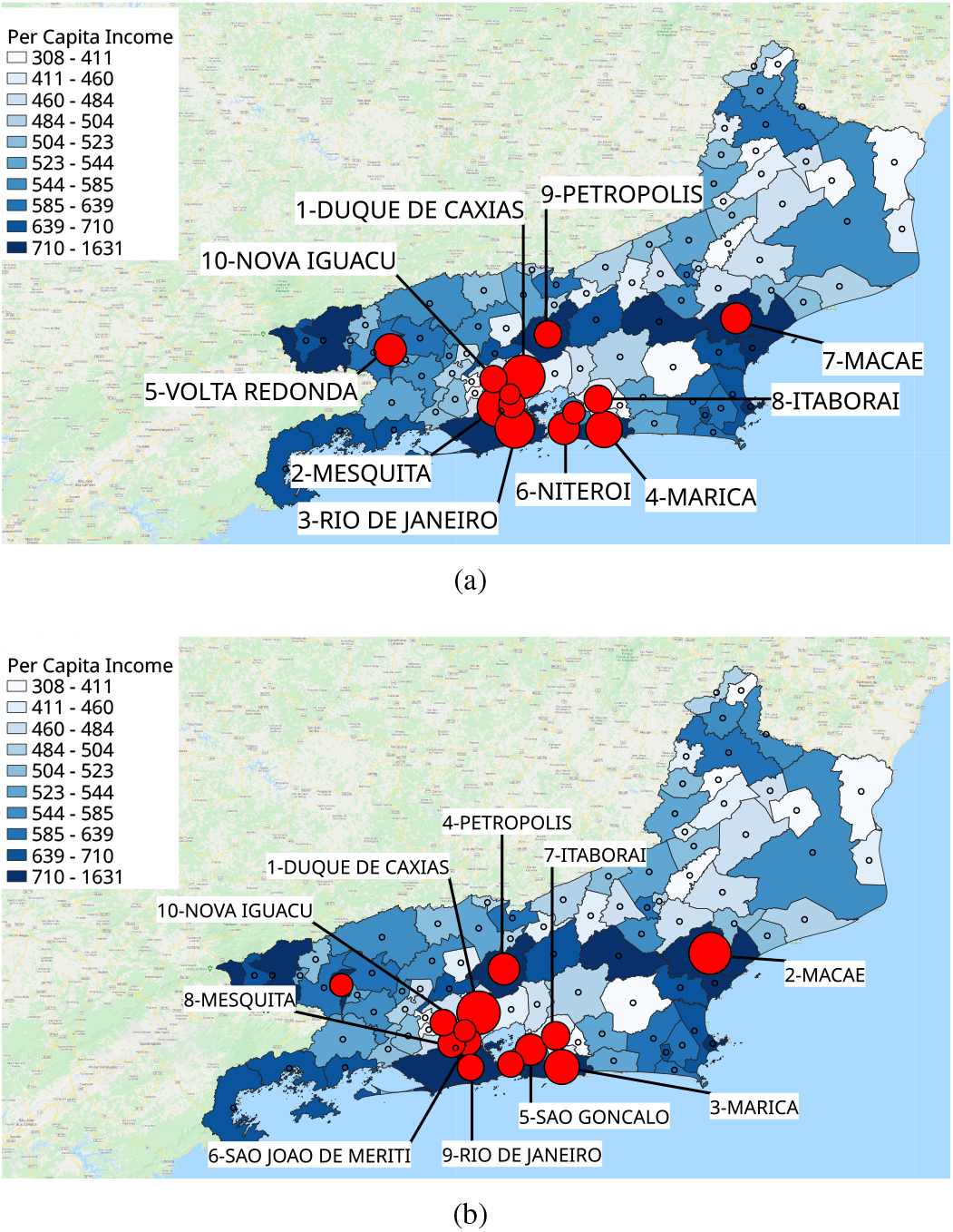
(a) Mortality and (b) lethality, for municipalities in the RJ state. The^10^ municipalities with the highest death rates in the state are presented. Empty circles represent locations with a number of deaths less than 7.

The lethality rate is shown in Fig. 8(b), where again the municipality of Duque de Caxias ranks first and Nova Iguaçu cupies the tenth. The rest of municipalities don’t remain in the same positions in relation to the mortality rate. However, an important observation to be made is that among the 10 municipalities with the highest lethality rates, 5 of them occupy the lower half of the per capita income stratum (Duque de Caxias, São Gonçalo, São João de Meriti, Itaboraí and Nova Iguaçu).

*\* Recovered*: The distribution map of the index of recovered people for every 100 thousand inhabitants is represented in Fig. 9(a). An interesting result on this map is that among municipalities belonging to the lower income quartiles per capita, only 3 of these municipalities appear among the 10 municipalities with higher recovery rates, occupying the positions numbers 5, 8 and 10. When considering the map of recovery rate by confirmed cases (Fig. 9(b)), we have the same situation. It is observed that these results are substantially different from those found for mortality and lethality rates.

**Figure 9:**
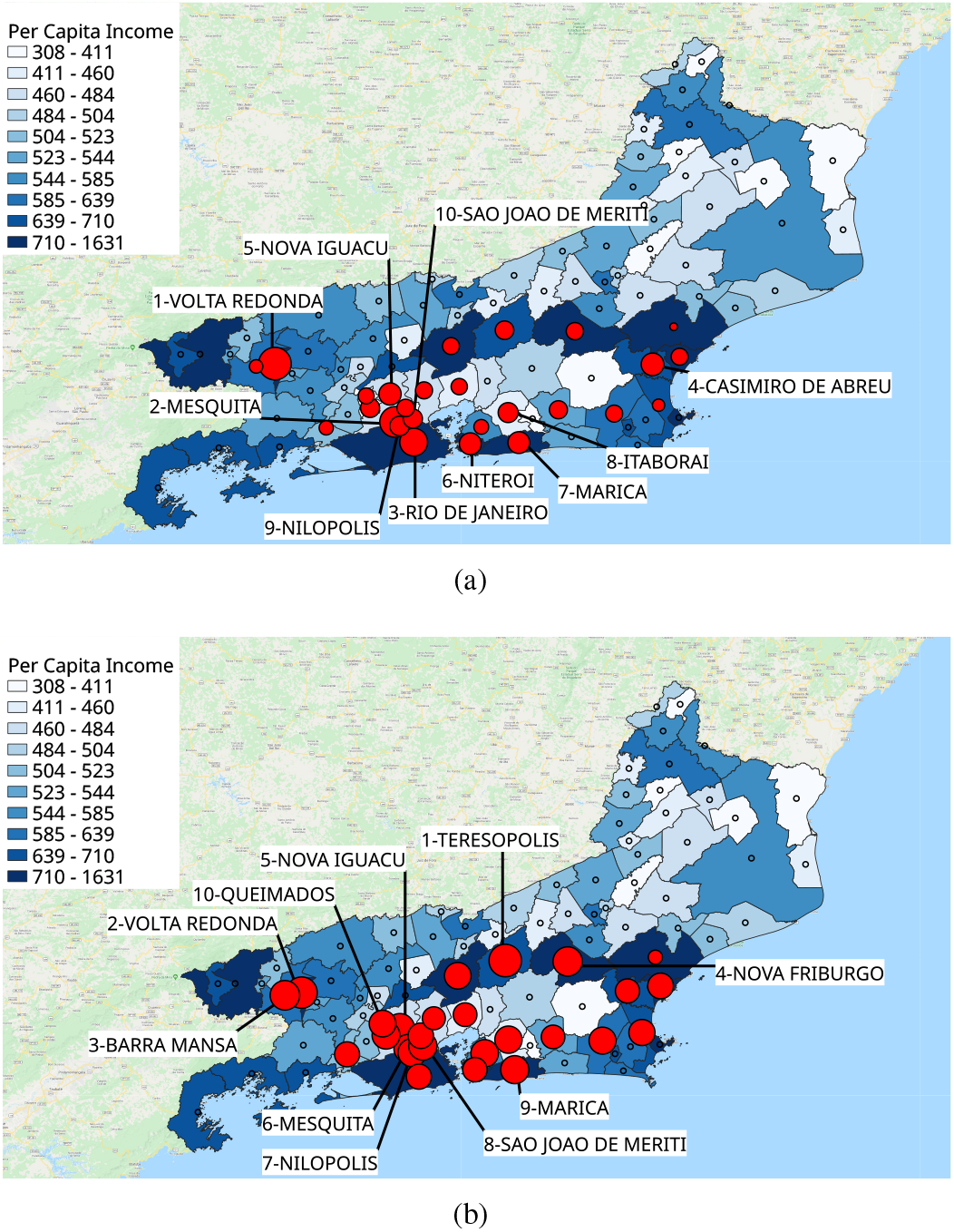
(a) People recovered, per 100 thousand inhabitants, for the municipalities of the state of RJ. (b) Ratio between the number of people recovered and confirmed cases for the municipalities of RJ. In both cases, empty circles represent locations with less than 20 confirmed cases of COVID-19. In both cases, the 10 municipalities with the highest rates of people recovered in the state, are highlighted on the map.

#### 5.2.2. Maps for neighborhoods in the municipality of Rio de Janeiro

*\* Confirmed cases*: The spatial distribution map of cases confirmed per 100,000 inhabitants in the municipality of Rio de Janeiro is shown in Fig. 10. It can be noted that the largest rates of confirmed cases are found in regions of high per capita income of the municipality of Rio, since among the 10 neighborhoods with the highest rates of confirmed cases, 9 have average per capita income belonging to the highest level of the map, with only 1 exception being the Bonsucesso neighborhood. An observation to be made is the Joá neighborhood, which despite being in the 5th position, had only 2 confirmed cases on the collection date of our data. However, because it has a very small value compared to the average, ends up being between the neighborhoods with higher rates of cases per population.

**Figure 10:**
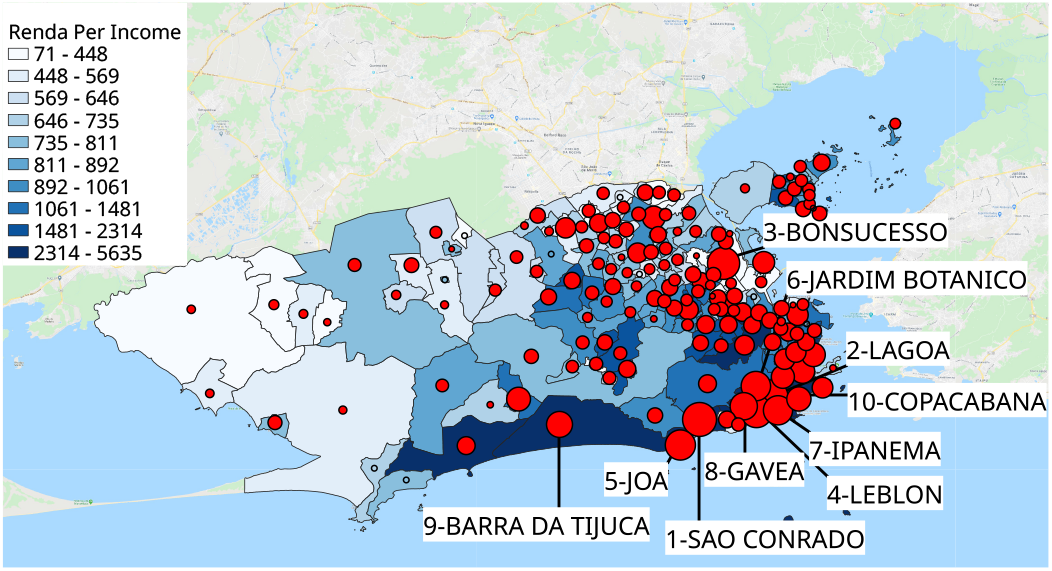
Confirmed cases for neighborhoods in the municipality of Rio de Janeiro. Empty circles represent locations with no confirmed case. Highlighted on the map are the 10 municipalities with the highest rates of confirmed cases in the municipality.

*\* Deaths*: As in the case of municipalities, we will use in our calculations only neighborhoods that have more than 6 deaths in our data.

The mortality rate is illustrated on the map in Fig. 11(a). In this figure we can see results different from those found for the rate of confirmed cases, given that for the mortality rate, only 6 high-income municipalities are among the 10 with the highest mortality.

**Figure 11:**
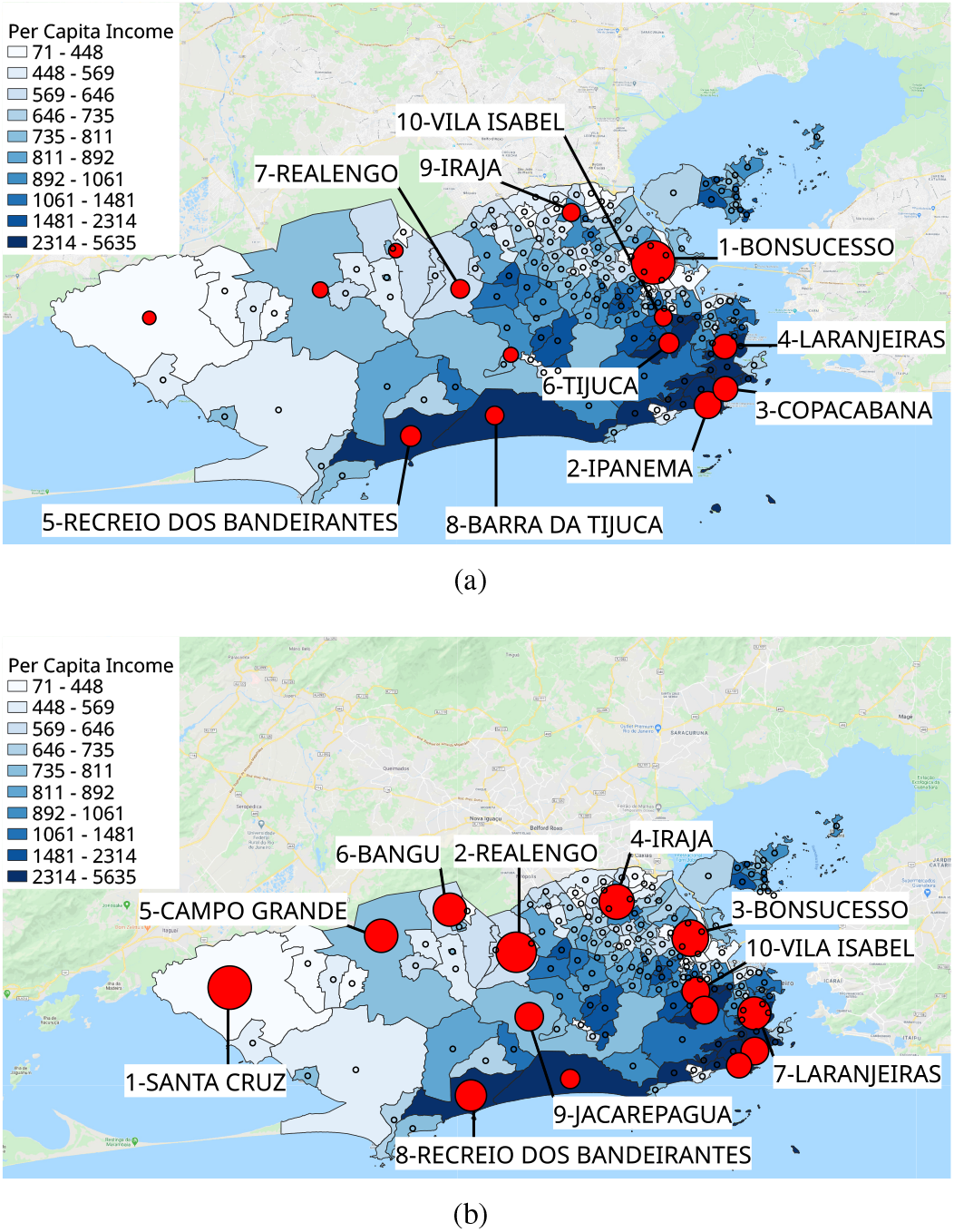
(a) Mortality and (b) lethality, for neighborhoods in the municipality of Rio de Janeiro. The 10 neighborhoods with the highest rates of deaths in the municipality are presented. Empty circles represent locations with an amount deaths below 7.

Fig.11(b) shows the lethality distribution map of neighborhoods in the municipality of Rio. For this map we have a situation where neighborhoods with high average per capita income correspond to only 2 of the most lethal neighborhoods, occupying the positions of numbers 7 and 8. It is also observed that the neighborhood that occupies the highest lethality position is in the municipality of Santa Cruz, where the average per capita income of population falls into the lowest income group.

In the maps of Fig. 11, similarly to what happened for the analysis of municipalities, there are dissonant results between the mortality-lethality rates from the results found for confirmed case rates.

*\* Recovered*: Fig. 12(a) represents the spatial distribution for the number of people recovered, per 100 thousand inhabitants, in the municipality of Rio de Janeiro. Following a behavior similar to that presented for the number of confirmed cases, 9 neighborhoods belonging to the highest per capita income group are among the 10 neighborhoods with the highest rate of people recovered. The exception is the Bonsucesso neighborhood. Yet, all 10 neighborhoods belong to the top half of the quartiles per capita income.

**Figure 12:**
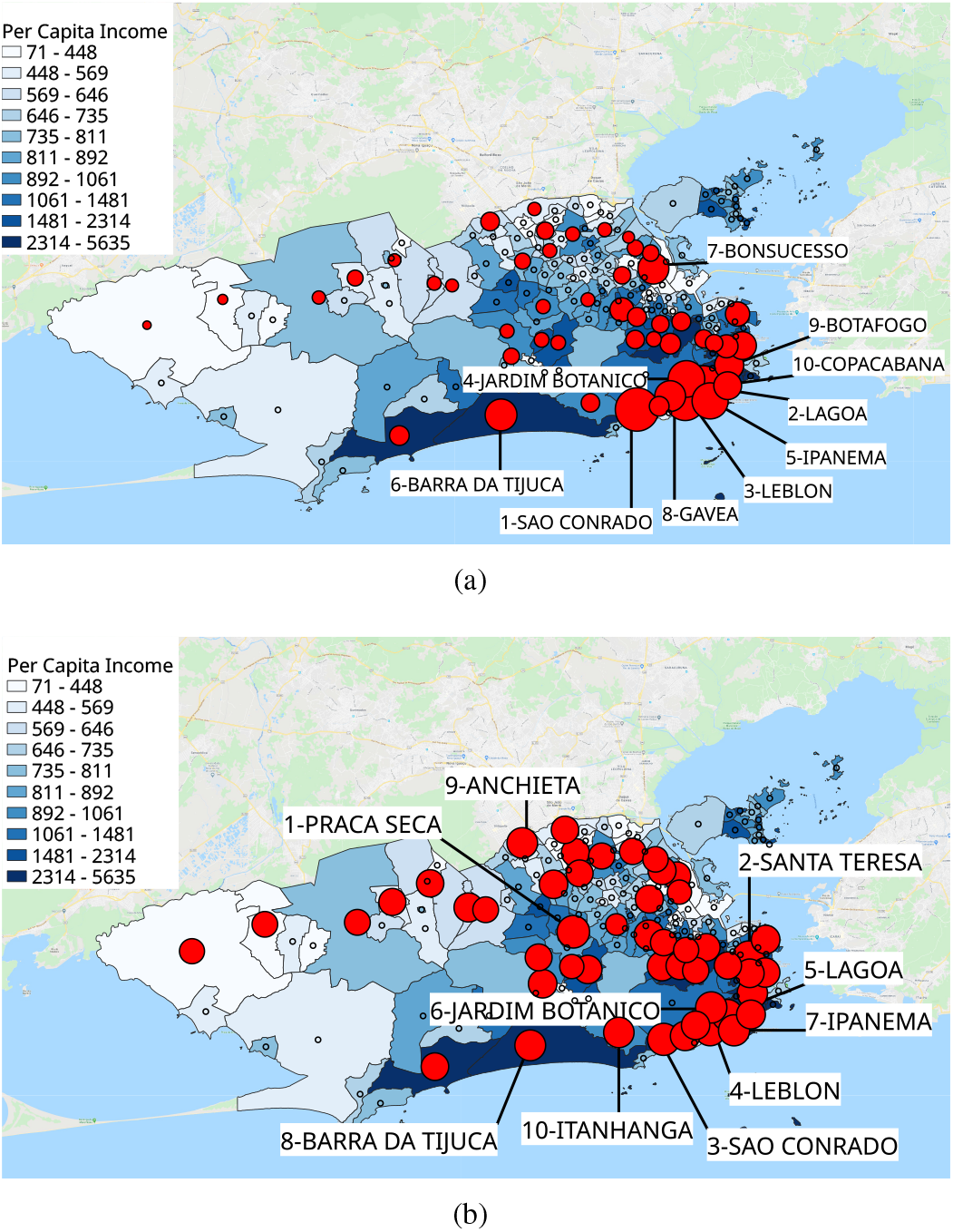
(a) People recovered, per 100 thousand inhabitants, for the neighborhoods in the municipality of Rio de Janeiro. (b) Ratio between the number of people recovered and confirmed cases for neighborhoods in the municipality of Rio de January. In both cases, empty circles represent locations with less than 20 confirmed cases of COVID-19. The 10 neighborhoods with the highest rates of people recovered in the municipality are presented.

The map of people recovered rate in relation to the number of confirmed cases is shown in Fig. 12(b). In this case the neighborhoods with the highest income per capita represent 6 of 10 neighborhoods with the highest recovery rate by confirmed cases. However, 9 of these 10 neighborhoods belong to the upper half of the per capita income distribution groups.

It is noted, therefore, that, as occurred for municipalities, these results are considerably different from those found for mortality and lethality rates.

### 5.3 Statistical analysis of spatial correlation

As described in section 4.3, one of the methods of producing a better understanding of the spread of a disease in a given region is the search for spatial correlations through quantitative methods.

We will present here results regarding the use of statistics Moran’s *I* applied to our epidemic and regional data. In order to check for local and regional correlations and the significance of the similar values found, we also apply in our data the local association method *LISA (Local Indicator of Spatial Association)*. For making maps and calculating the respective indexes presented in this section, the GeoDa software was used^11^.

Similar to what was done for the space distribution study, in section 5.2, we will divide the statistical analyzes of spatial correlation at two levels of spatial aggregation: for the municipalities of the state of RJ and for the neighborhoods in the municipality of Rio de Janeiro.

#### 5.3.1. Statistical analysis for the municipalities of the state of RJ

Since Moran’s *I* statistic needs the distribution of weights assigned between each region and the neighbors considered for this region, we present in Fig. 13 the histogram of distribution of numbers of neighbors for the municipalities of the RJ state. To calculate this histogram we used the neighborhood form with *queen*-type contiguity criteria [35]. This histogram provides a measure of the number of neighbors that each municipality has.

**Figure 13:**
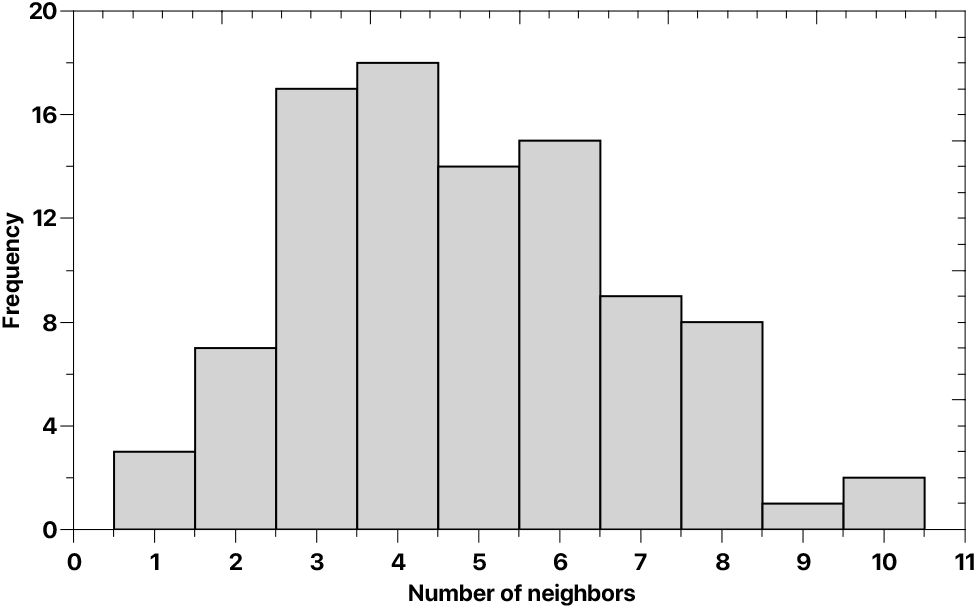
Distribution histogram of the number of neighbors to the municipalities of the state of RJ.

The statistical analyzes will be presented below using the division by: confirmed cases, deaths and recovered.

*\* Confirmed cases*: For the municipalities of the state of RJ the Moran’s *I* presented a value equal to 0.371, indicating the existence of significant spatial autocorrelation between municipalities. In view of the positive value of index *I*, we have a direct correlation between regions, that is, we can observe that locations with high rates of confirmed cases tend to be close to other locations that also have high rates and low-rate locations tend to be close to neighborhoods with low rates too. This result is evidenced in the dispersion graph of Moran’s index *I* (Fig. 14(a)), with predominance of points in quadrants I and III.

**Figure 14:**
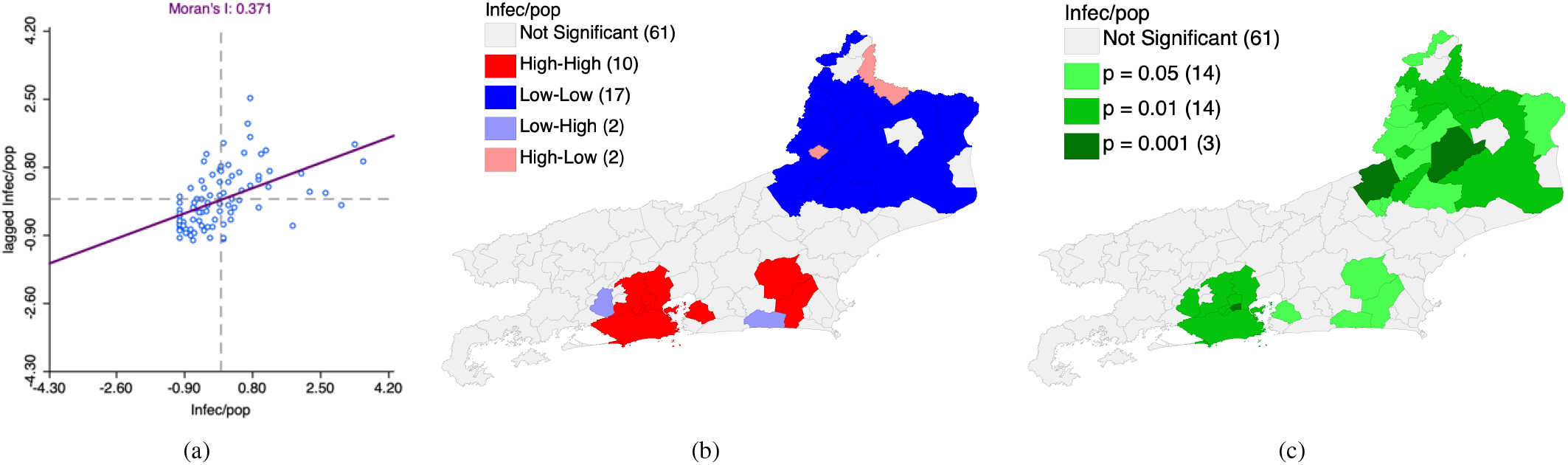
*Moran’s I statistics* applied to confirmed cases of COVID-19 in the cities of the state of RJ. (a) Scatterplot for Moran’s index *I* for the rate of confirmed cases of infected persons for a population of 100 thousand inhabitants. (c) Map of clusters – LISA. (b) Map of significance –LISA.

Applying the LISA method, autocorrelations were observed with the presence of spatial clusters and significant statistic. The cluster map and the significance map shown in Figs. 14(b) and 14(c) expose these results.

The maps indicate the formation of a large cluster of confirmed casualties rates for COVID-19 in upstate and a high-rate cluster in the region close to the greater Rio. However, we can see that the municipalities of Seropédica and Saquarema, are found in *low-high* regions, that is, despite having low rates of confirmed cases, they are close to municipalities with high rates, making that the risk of growth in the number of cases in these regions become higher. Conversely, we have the municipalities of Aperibé and Bom Jesus do Itabapuana presenting themselves as regions *high-low* type, since they have high case rates confirmed, but their neighbors have low rates.

*\* Deaths*: Moran’s index *I* for death rate, due to the coronavirus, in the municipalities of the state of RJ, did not indicate the existence of spatial autocorrelation, since the value found *I* = 0.059 indicates a spatial independence, given that the value is close to zero. Likewise, the map of clusters – LISA did not identify the presence of autocorrelations between municipalities.

As no relevant statistical results were identified for the spatial correlations between municipalities, the graph of dispersion for Moran’s *I* index and LISA maps will not be presented here.

*\* Recovered*: Similar to cases of death, there is not the presence of spatial correlations between the municipalities in Rio de Janeiro regarding the recovery rates of people infected with COVID-19. In this case, the index *I* of Moran presented result *I* = −0.014, where again we have the indicative of spatial independence. The cluster map – LISA also doesn’t identify the presence of autocorrelations between municipalities for recovered people.

#### 5.3.2. Statistical analysis for neighborhoods in the municipality of Rio de Janeiro

For Moran’s *I* statistical analysis applied to the dissemination of COVID-19 in the neighborhoods in the city of Rio de Janeiro, we initially present the distribution of the numbers of neighbors for the neighborhoods. The histogram shown in Fig. 15 was calculated using the *queen* contiguity criterion for the neighborhood shape [35].

**Figure 15:**
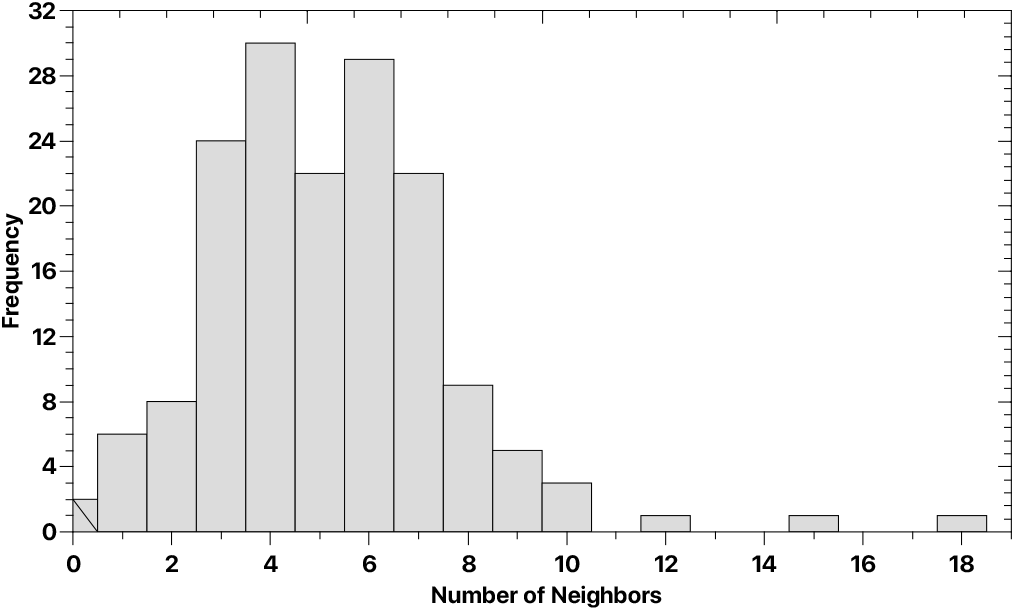
Distribution histogram of the number of neighbors for the neighborhoods in the municipality of Rio de Janeiro.

One observation to be done in this statistical analysis is that the municipality of Rio de Janeiro has two neighborhoods that do not have neighbors (see Fig. 15). In this way, these two neighborhoods are excluded when performing the statistical calculations performed in the following steps.

*\* Confirmed cases*: The spatial distribution of confirmed cases of COVID-19 in the neighborhoods of the municipality of Rio de Janeiro presented *I* = 0.484 for Moran’s *I* index, with the most of the variances in relation to the average distributed in quadrants I and III, as shown in the scatter plot for Moran’s *I* index, in Fig. 16(a). This result indicates the existence of significant spatial autocorrelation between neighborhoods in Rio de Janeiro regarding confirmed cases of COVID-19. Similar to what happened for municipalities of the state of RJ, as we have *I* > 0, the neighborhoods have a direct spatial correlation, in which regions with high rates of cases tend to have neighborhoods that also have a high rate, and neighborhoods with low rates tend to be close to other regions with low rates.

**Figure 16:**
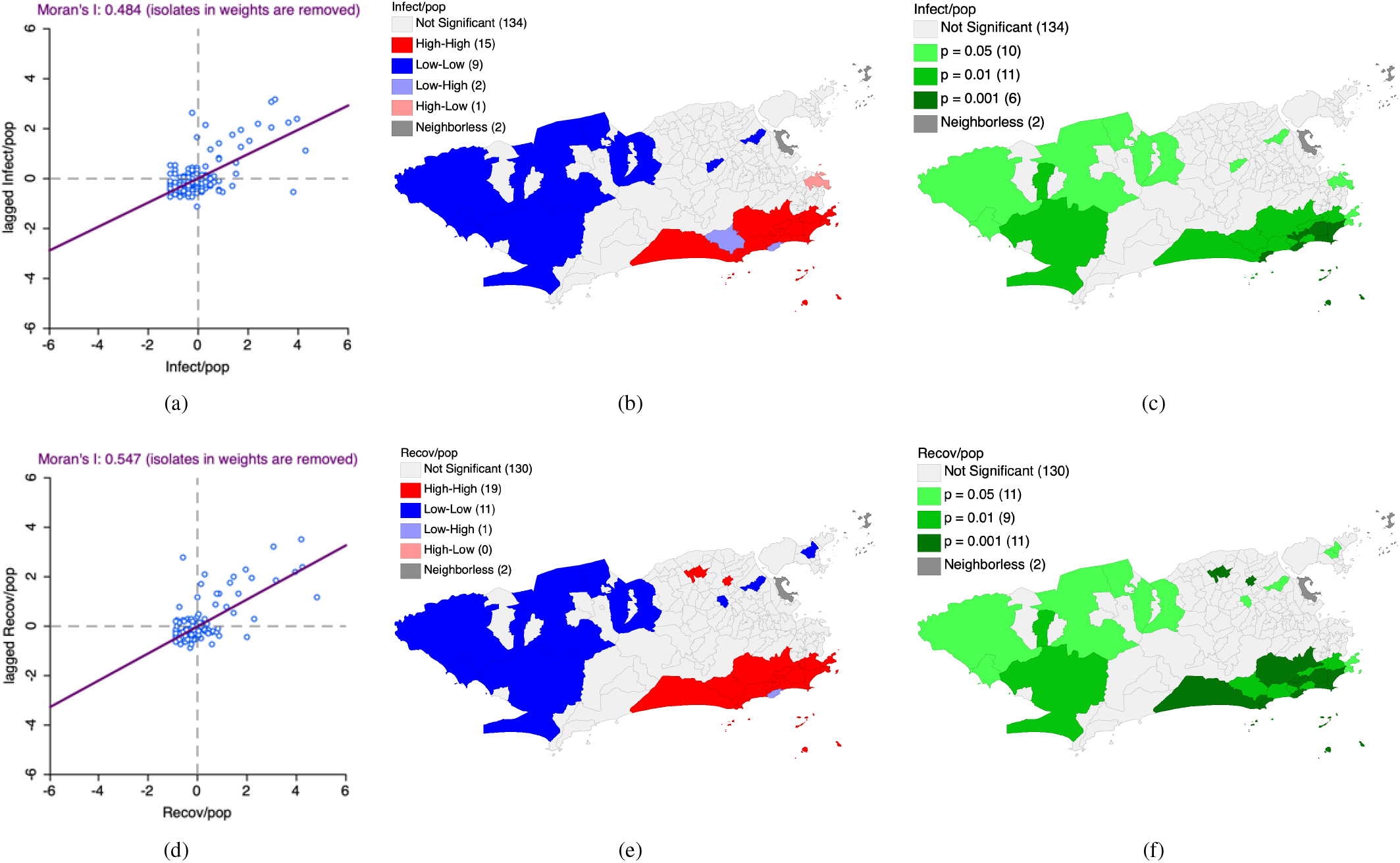
*Moran’s I statistics* applied to COVID-19 data for neighborhoods in the municipality of Rio de Janeiro. (a) Scatterplot for index *I* of Moran for the rate of confirmed cases of infected people for a population of 100 thousand inhabitants. (b) Map of clusters – LISA for the rate of confirmed cases. (c) Map of significance – LISA the rate of confirmed cases. (d) Scatterplots for Moran index *I* for the rate of people recovered for population of 100 thousand inhabitants. (e) Map of clusters – LISA for the rate of people recovered. (f) Significance map – LISA for the rate of people recovered.

The LISA method applied to confirmed cases for neighborhoods of Rio provides the maps shown in Figs. 16(b) and 16(c), where the presence of spatial clusters and significance results is observed.

Figs. 16(b) and 16(c) show a large cluster of neighborhoods with high rate of confirmed cases of COVID-19 (*high-high*) located in the region that has the highest income per capita neighborhoods of the municipality. In addition, a large cluster of low rate (*low-low*) regions is located in the western of the municipality. The exceptions are the neighborhoods of Olaria and Cascadura, which also present themselves as regions *low-low* type, but are located outside the western zone.

When observing the neighborhoods of Itanhangá and Vidigal, one can notice that both have low rates of confirmed cases, however, they are surrounded by neighborhoods with a high case rate, and therefore present themselves as *low-high* regions. These two neighborhoods do not fall into the category of neighborhoods with higher average per capita incomes. The Centro neighborhood is the only one present in the *high-low* category, that is, it has a high rate of cases confirmed for COVID-19, immersed in a region where its neighbors have low rates.

It should be noted that a result similar to this one, of cases confirmed for the neighborhoods of Rio de Janeiro, was found in [6], using data between March 6, 2020 and April 10, 2020.

*\* Deaths*: Similar to what happened for state municipalities, the death rate for neighborhoods in the municipality of Rio de Janeiro showed no spatial correlation, considering that the Moran index *I* obtained was equal to −0.007, indicating therefore, a spatial independence between the neighborhoods.

Also, the cluster map for the death rate of Rio’s neighborhoods did not identify a significant amount of clusters as to indicate the presence of spatial autocorrelations between the neighborhoods. With that, the scatterplot for index *I* Moran and LISA maps will not be shown here.

*\* Recovered*: Results for the rate of recovered people are similar to those found for confirmed cases. The Moran index I found was 0.547, indicating also the existence of significant spatial autocorrelation between the neighborhoods of Rio de Janeiro with regard to people recovered of COVID-19 infection. The concentration of points in quadrants I and III of Fig. 16(d) again reflects the correlation between neighborhoods in relation to the rate of recovered.

Similar to confirmed cases, the LISA method presented a large *high-high* cluster in the region of higher per capita income (maintaining the Vidigal neighborhood as a region *low-high*) and another large *low-low* cluster in the area west (with Olaria also remaining in a region of this type). However, for the rate of recovered, the neighborhoods of Coelho Neto, Barros Filho and Itanhangá present themselves as zones *high-high* and the neighborhoods of Cavalcanti, Jardim Carioca and Tauá as *low-low* regions. These results are presented on the maps of Figs. 16(e) and 16(f).

In order to obtain quantitative results regarding the existence of spatial correlations between the rate of confirmed cases and the average per capita income of the neighborhoods, we perform the bivariate Moran *I* statistics, using these parameters as two income variables, cases and income. The same procedure was done for the rate of people recovered in relation to neighborhoods per capita income. The results are shown in Figs. 17(a) e 17(b), where it is possible to observe that Moran’s index *I* is equal to 0.529 for the COVID-19 confirmed case rate and equal to 0.573 for the rate of recovered people, indicating direct (positive) spatial correlation between these rates and average income per capita of the neighborhoods, thus confirming what was previously presented, that is, for the data considered in the present study, the neighborhoods with the highest per capita income are correlated to greater numbers of cases and people recovered, while the lower middle income neighborhoods are correlated to fewer cases and people recovered.

**Figure 17:**
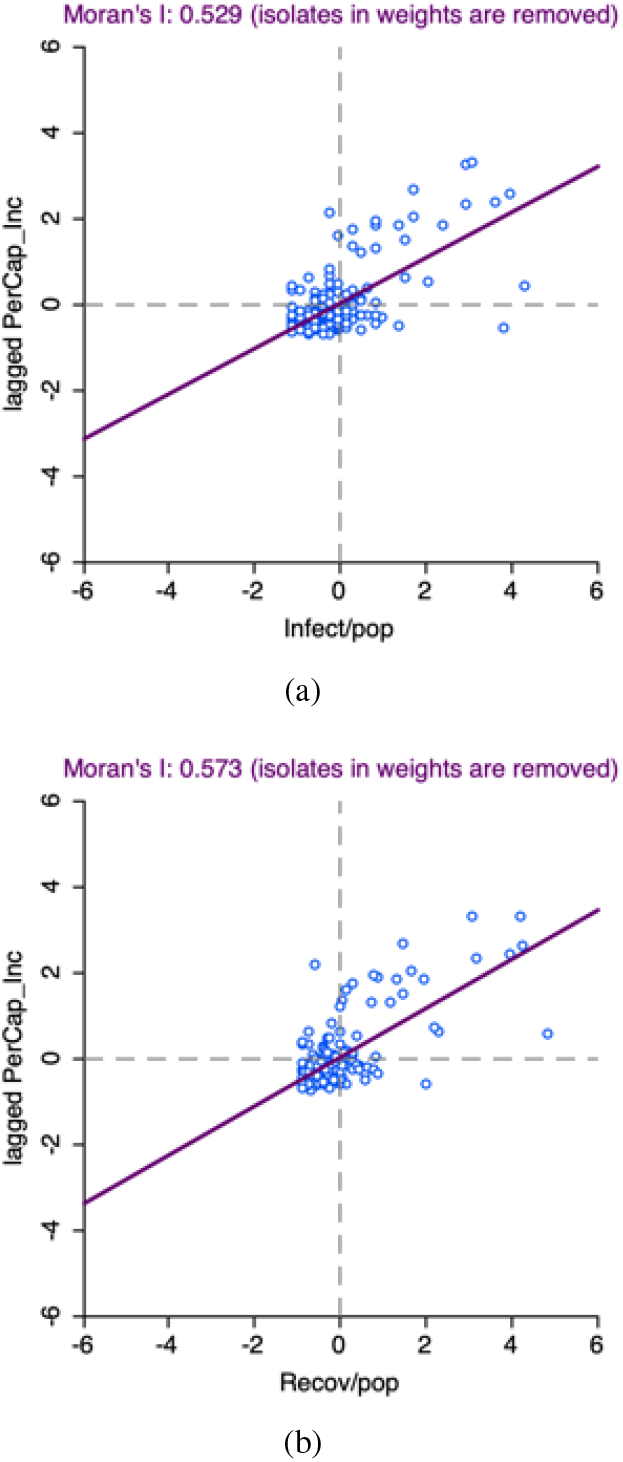
Bivariate *Moran’s I statistics* applied to COVID-19 data for neighborhoods in the municipality of Rio de Janeiro. Scatterplots for the Moran index *I*(a) for the rate of confirmed cases of infected people for population of 100 thousand inhabitants and (b) for the rate of recovered people.

## 6. Conclusions

In the present work, we first analyzed the evolution of confirmed cases of COVID-19, death rates and rates of recovered people, where death rates were divided into mortality and lethality rates, and the rates of recovered people were calculated per 100 thousand inhabitants and for confirmed cases. These analyzes were made under three levels of regional scales, namely, for the entire state, for the municipalities and neighborhoods in the municipality of Rio de Janeiro. Our results point to an exponential behavior, in the early stages of dissemination, of rates of confirmed cases, of mortality and also for recoveries, at all levels of scale (state, municipalities and neighborhoods). It is worth mentioning that these results corroborate studies about the dissemination of COVID-19 in Brazil and other regions of the world, as well as for other contagious diseases [17, 18, 19, 20, 21, 22, 23, 24].

In addition, it was observed that some municipalities such as Duque Caxias and Rio de Janeiro have mortality rates considerably higher than the other municipalities, with Duque de Caxias still having a high lethality rate, which behaves in an approximately as a constant from the 30th day on. At the same time, the municipality of Niterói, for a time span of 15 days, has a rate of recovery cases markedly higher than other municipalities. Considering an exponential spread, such as for COVID-19, an interval of 15 days in which there are high recovery rates, can mean many lives saved.

The second method adopted uses a spatial analysis to map the distribution of the COVID-19 outbreak by municipalities of the state of RJ and the neighborhoods of the municipality of Rio de Janeiro. With that it was possible to observe a considerably incidence of lower number of cases in the north of the state. Another result obtained was that in our analysis Duque de Caxias presents itself as the municipality with the highest mortality and lethality in the state of Rio de Janeiro.

Regarding the municipality of Rio de Janeiro, it was observed a strong trend of confirmed cases and recovery rates in the south of the municipality. At the same time, for the lethality rate, among the 10 largest neighborhoods, only 1 belongs to the southern part of the municipality.

The last method used was the analysis performed through the Moran’s *I* statistics in order to obtain spatial correlations between the municipalities of the state and between neighborhoods in the capital of RJ. For the municipalities, a significant direct spatial autocorrelation was observed, in which it is confirmed that northern municipalities tend to be cluster low rates of cases of COVID-19 and the municipalities close to the Greater Rio tend to form high-rate clusters. For neighborhoods in the municipality of Rio, a strong space autocorrelation was also found, in which the municipalities of the southern part of the state concentrate most of the high-rate cluster, while the western zone concentrates most of the neighborhoods of low rates.

Although the three used methods are independent, it is observed that their results are reinforced and complemented by each other. It can be highlighted, for example, that all three methods point to the fact that the higher average income per capita regions tend to be places with a higher number of cases confirmed COVID-19 and recovered persons. This result was also confirmed for the fourth time with the use of Moran’s bivariate *I* statistic pointing to a strong spatial correlation between locations with higher case rates and recovery and neighborhoods in the municipality of Rio de Janeiro with higher per capita income.

New studies to assess future trends of rates of cases, deaths and recoveries for municipalities and neighborhoods can be direct consequences of this article. We expect the present work to contribute in the analyzes of creating specific strategic plans in municipalities (and even in the neighborhoods) for an effective and efficient fight against spread of the new coronavirus.

## Data Availability

All data referred to in the manuscript are availability in Health Departments of Governments of the State of RJ and the Municipality of Rio de Janeiro.

## Acknowledgements

J.R. thanks for the scholarship of FAPERJ, Brazil in this research, P.S.L.O thanks for the scholarship of CNPq, Brazil, A.R.R.P thanks CNPq, Brazil, for productivity grant and D.S.R.F thanks FAPERJ, Brazil, for grant number 241029.

## Footnotes

1 https://www.who.int/news-room/detail/27-04-2020-who-timeline-covid-19

2 https://covid.saude.gov.br

3 http://painel.saude.rj.gov.br/monitoramento/covid19.html

4 http://www.prefeitura.rio/coronavirus

5 https://cidades.ibge.gov.br/brasil/rj/panorama

6 https://cidades.ibge.gov.br/brasil/rj/rio-de-janeiro/panorama

7 https://cps.fgv.br/r-renda-capita-populacao-total-e-favelas-municipios-dorio-de-janeiro

8 https://cps.fgv.br/r-renda-capita-populacao-total-e-favelas-bairros-rio-dejaneiro

9 http://covid.saude.gov.br

10 https://www.qgis.org/en/site/

11 https://geodacenter.github.io

